# Identifying Factors Associated with Lower Quarter Performance-Based Balance and Strength Tests: the Project Baseline Health Study

**DOI:** 10.1101/2023.08.23.23294526

**Authors:** Kenneth A. Taylor, Megan K. Carroll, Sarah A. Short, Bettia E. Celestin, Adam Gilbertson, Christoph B. Olivier, Francois Haddad, Nicholas Cauwenberghs

## Abstract

**Background:** Physical performance tests are predictive of mortality and have been proposed for screening for certain health conditions (e.g., sarcopenia); however, the diagnostic screening and prognostic value of physical performance tests has primarily been studied in age-limited or disease-specific cohorts. In this study, we sought to identify the most salient characteristics associated with three lower quarter balance and strength tests in a deeply phenotyped cohort of community-dwelling adults.

**Methods:** We applied a stacked elastic net approach on detailed data on sociodemographic, health and health-related behaviors, and biomarker data from the first visit of the Project Baseline Health Study (N=2502) to determine which variables were most associated with three physical performance measures: single-legged balance test (SLBT), sitting-rising test (SRT), and 30-second chair-stand test (30CST). Analyses were stratified by age (<65 and ≥65).

**Results:** Female sex, Black or African American race, lower educational attainment, and health conditions such as non-alcoholic fatty liver disease and cardiovascular conditions (e.g., hypertension) were consistently associated with worse performance across all three tests. Several other health conditions were associated with either better or worse test performance, depending on age group and test. C-reactive protein was the only laboratory value associated with performance across age and test groups with some consistency.

**Conclusions:** Our results highlighted previously identified and several novel salient factors associated with performance on the SLBT, SRT, and 30CST. Future research should discern and validate the value of these tests as affordable, noninvasive biomarkers of prevalent and/or future disease in the community.

## INTRODUCTION

A person’s ability to perform activities of daily living - physical function - can worsen by disease or injury of the neurological, cardiopulmonary, musculoskeletal or other physiological systems and is affected by other clinical and sociodemographic factors associated with unfavorable health-related behavior.^1^ Declines in physical function are often subtle and unnoticed until they reach a stage at which preventive measures may be less effective, making it important to identify people with subclinical declines in physical function to initiate effective preventive interventions early.^2–5^

For this, standardized clinical tests have been developed, which enable affordable and objective quantification of different aspects of physical function, including control of static posture and balance, musculoskeletal fitness and strength, and overall mobility. Performance on several physical function tests have been identified as health status indicators of prevalent health conditions and predictive of downstream health outcomes and all-cause mortality, particularly in older individuals.^6–15^

To date, these physical performance tests have primarily been studied in age-limited or diagnosis-specific cohorts to assess their relationships with specific clinical characteristics. Such studies are useful, but have limited ability to identify potential relationships between physical performance tests and a wider range of prevalent sociodemographic and health characteristics.^16^ Sociodemographic and clinical correlates of physical function tests have yet to be assessed in a more comprehensive way. Data from deeply phenotyped cohorts could facilitate the identification of novel associations between physical function tests and characteristics that warrant further investigation. Therefore, we aimed to identify the most salient characteristics associated with the three lower quarter physical performance tests (single-legged balance [SLBT], sitting-rising [SRT], and 30-second chair-stand [30CST]), from sociodemographic and health features as recorded in a deeply phenotyped cohort of community-dwelling adults, stratified by age (≥65 years-old and <65 years-old).

## METHODS

### Project Baseline Health Study

In this study, we used data from The Project Baseline Health Study (PBHS). The PBHS is a multicenter longitudinal cohort study of adults in the United States (ClinicalTrials.gov identifier NCT03154346), approved by both a central Institutional Review Board (the WCG IRB; approval tracking number 20170163, work order number 1-1506365-1) and IRBs at each of the participating institutions: Stanford University (Palo Alto, CA), Duke University (Durham, NC; Kannapolis, NC), and the California Health and Longevity Institute (CHLI; Los Angeles, CA). Participants are deeply phenotyped and contribute data including demographic characteristics, socioeconomic status-related health behaviors, medical conditions, symptoms, and laboratory-collected biomarkers in addition to physical function markers and a range of patient-reported outcome measures. A full description of study procedures and design, inclusion and exclusion criteria, and institutional review board approval has been previously made available.^17,18^

This study used cross-sectional data from the first in-person study collection time point; self-reported data used were collected at either the same in-person timepoint or via remote application. We limited data that were only collected remotely to participant responses within 200 days of participants’ initial in-person data collection.

### Physical Performance Measures

#### SLBT

It assesses static postural and balance control with moderate-to-excellent test-retest reliability and excellent inter-rater reliability.^19,20^ To perform the SLBT, participants were instructed to place their hands on their hips with their eyes open while focusing on a point straight ahead and stand on one leg while raising the other. The time was recorded from when one foot was lifted from the floor until whichever occurred first: the lifted foot touched the stance leg or the ground, the stance foot moved on the floor, either hand left the hips, or 60 seconds had passed. The test was performed on each leg one time and the SLBT time recorded was the mean of the left and right leg balance durations. Poor performance on the SLBT may help in predicting falls^21,22^ and inability to achieve at least 10 seconds has been recently linked to all-cause mortality.^14^

#### SRT

The SRT was developed to assess muscle strength and power, flexibility, and balance components of physical fitness through evaluating an individual’s ability to transfer from standing to sitting on the floor and then rise from the floor back to standing.^23^ The test has also been shown to predict all-cause mortality in a cohort of adults aged 51-80 years-old.^15^ To perform the SRT, participants were placed on a non-slippery flat surface in a minimum space of 2×2 meters without wearing shoes. The basic movements of the test were explained to the participant before the participants were instructed, “without worrying about the speed of movement, try to sit then to rise from the floor using the minimum support that you believe is needed.” The test was scored based on performance, with a maximum of 10 points - 5 points for transferring from standing to sitting on floor and 5 points transferring from sitting on the floor back to standing. Each participant started with a score of 5 for each movement from which points were deducted - 1.0 point deducted for each time a hand, forearm, knee, side of the leg (but not the sides of the feet), or hand on the knee was used and 0.5 point deducted for each time a partial loss of balance was observed. Participants who did not obtain a full score on either the sitting or rising portions of the test were provided additional advice or demonstration of the action by site staff and allowed to perform additional attempts to improve their scores. Regardless of the number of attempts performed, only the best scores for each sitting and rising attempt were recorded for the participant’s SRT score. Despite the rater-based scoring, high inter-rater reliability has been reported for the SRT.^24^

#### 30CST

This is a measure of functional lower extremity strength with excellent criterion, interrater, and test-retest reliability.^25^ To perform the 30CST, participants were positioned in a 17-inch high straight-backed armless chair and instructed to keep their feet flat on the floor and arms crossed over their chest during the test. Upon hearing “go” participants were instructed to rise to a full standing position and then back down to sitting for 30 seconds. The test is scored by recording the number of full stands completed in 30 seconds. Being >50% to a full standing position at the end of the 30 seconds was counted as a full stand. Since the test is scored using stand count, the 30CST does not have the same floor effect as other standing tests that are scored by recording time-to-completion for a specific number of stands (e.g., 5-time sit-to-stand).^26^

### Other Variables

Data collected from PHBS participants has been previously reported.^17^ Variables were considered for inclusion in regression analyses when age-specific prevalence was greater than 1%.^27^ In participants aged <65 years, this removed 13 medical conditions from the list of eligible candidate variables: atrial fibrillation, benign prostate hyperplasia, prostate cancer, breast cancer, hepatitis B, myocardial infarction, macular degeneration, melanoma, pulmonary embolism/deep vein thrombosis, peripheral vascular disease, stroke, transient ischemic attack, and goiter. In participants aged ≥65 years, we removed 6 conditions from the list of eligible candidate variables that had prevalence of <1%: bipolar disorder, type I diabetes mellitus, drug abuse, fibromyalgia, hepatitis C, and posttraumatic stress disorder (PTSD). We applied Box-Cox transformations to each laboratory value to approximate Gaussian distributions. We present an ascertainment and definition summary of eligible variables used in our analyses in the **Supplemental Material**.

### Statistical Analysis

We summarized clinical and demographic characteristics for participants with completed physical performance tests. All analyses were stratified by age and models were estimated separately for younger (<65 years) and older (≥65 years) participants. The SLBT was dichotomized for adults aged <65 years as either achieving the maximum time (60 seconds) or not (<60 seconds) based on the high proportion of individuals in this subgroup hitting the 60-second ceiling. SLBT was kept as continuous for adults aged ≥65 years-old, which had a wider distribution in this subgroup. The SRT and 30CST did not demonstrate similar ceiling effects and were left as continuous variables across both age subgroups. We used multiple imputation by chained equations to estimate values for missing data and fit regression models using elastic net (ENET) regularization methods. We employed ENET using a stacked objective function (sENET) with 5-fold cross-validation to penalize and select regression coefficients modeling each physical performance measure.^28,29^ A more detailed description of the sENET methods we used can be found in the **Supplemental Material**. We performed a sensitivity analysis that added self-reported symptoms to the list of eligible covariate candidates. Additional details and reasoning are presented in the **Supplemental Material**.

## RESULTS

### Participants

Of the 2502 participants in the PBHS cohort, 2339 (93.5%), 2198 (87.9%), and 2410 (96.3%) completed SLBT, SRT, and 30CST at the initial visit, respectively. Among those who completed physical performance tests, 23.6% (SLBT), 22.2% (SRT), and 23.0% (30CST) were aged ≥65 years. We present the age-stratified distributions and demographic characteristics by physical performance test in **Figure 1** and **Table 1**, respectively. For participants aged <65 years, mean (SD) physical performance test scores were 47 (20) seconds for SLBT, 7.7 (2.1) points for SRT, and 15 (5.2) stands for 30CST. For participants ≥65 years of age, mean (SD) test scores were 23 (20) seconds, 5.4 (2.3) points, and 14 (5.0) stands for SLBT, SRT, and 30CST, respectively.

**Figure 1.**
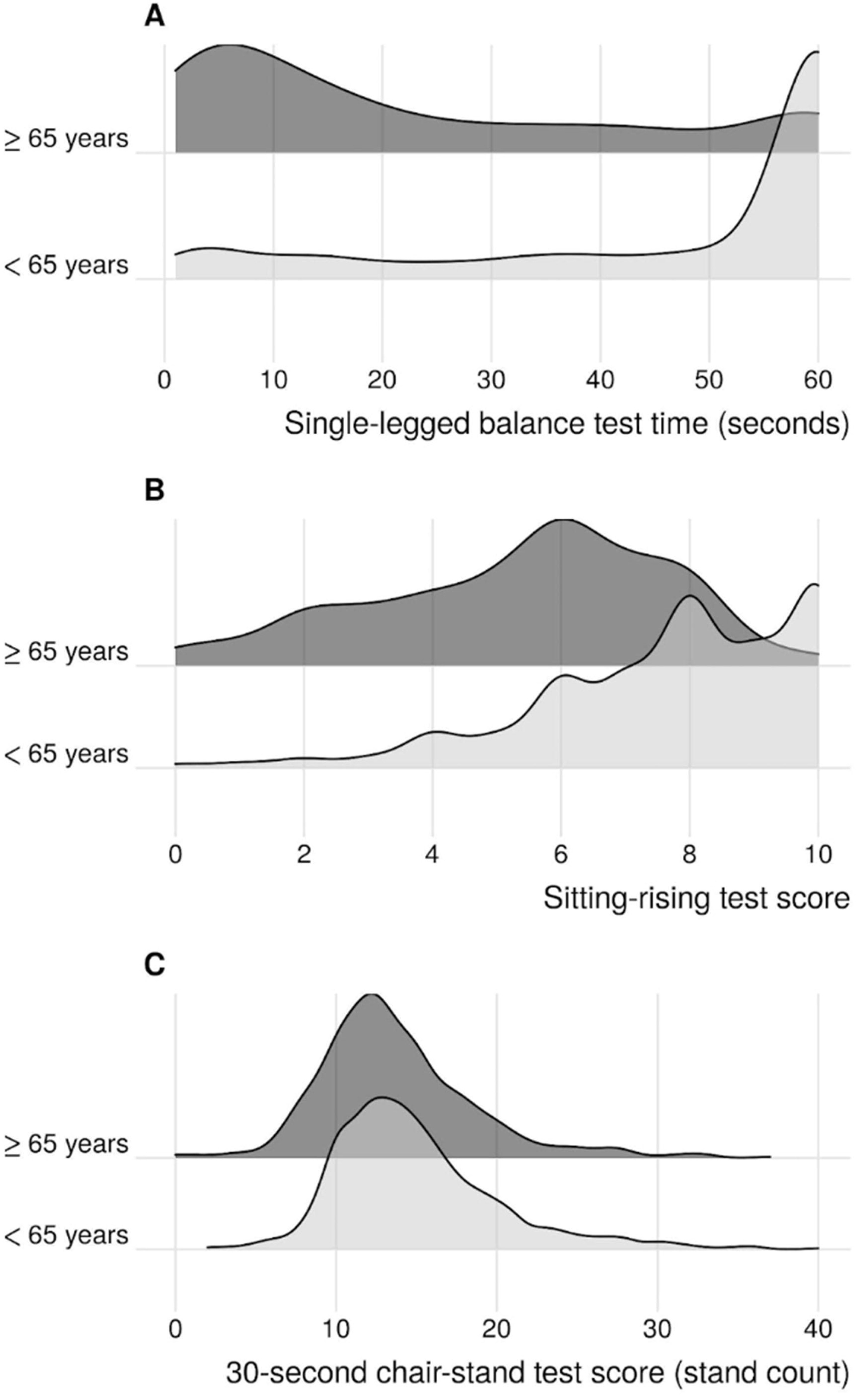
**Kernel density ridgeline plots of age-stratified physical performance results.** Panel A: Single-legged balance test; Panel B: Sitting-rising test; Panel C: 30-second chair-stand test.

**Table 1.**
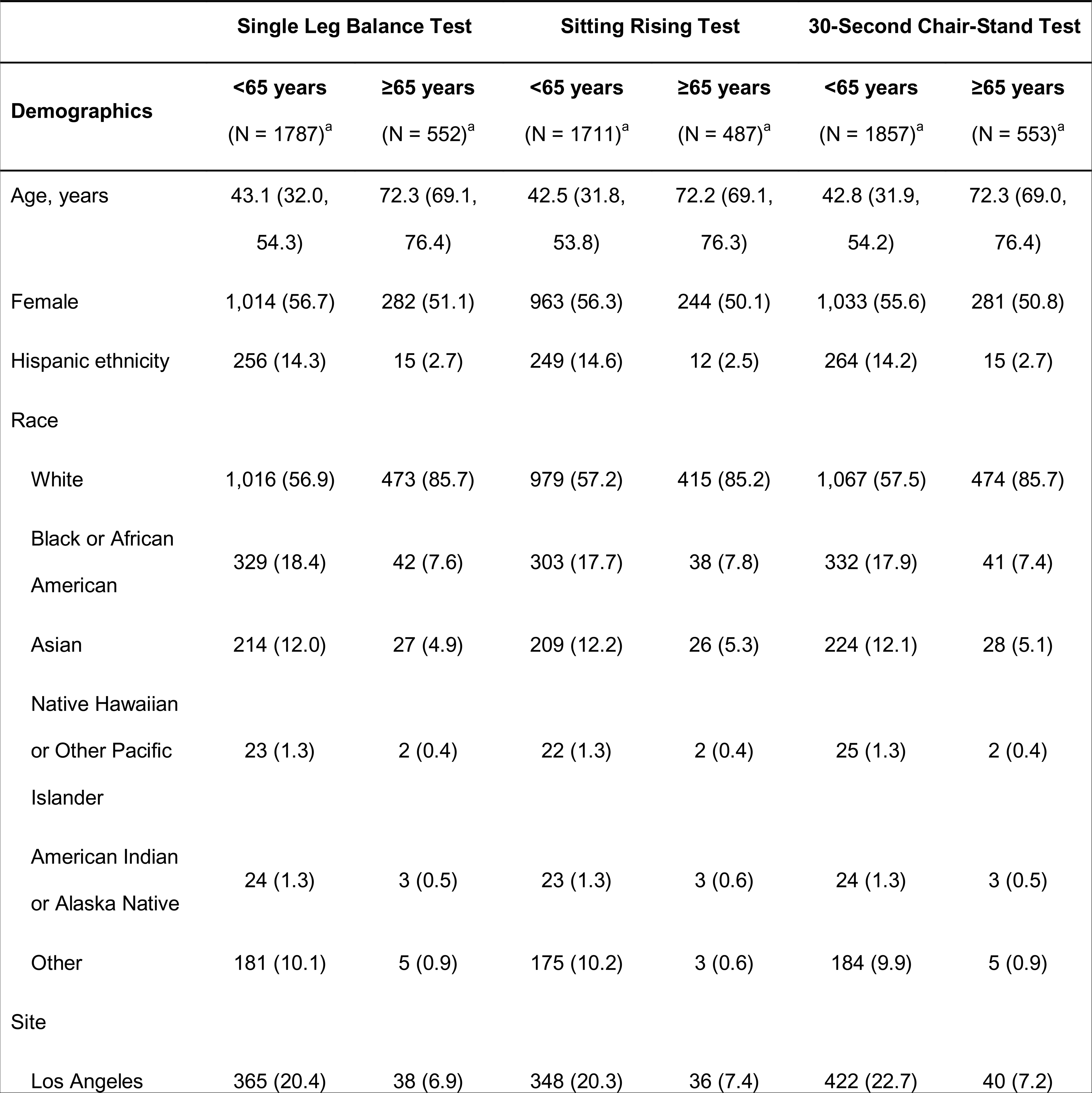
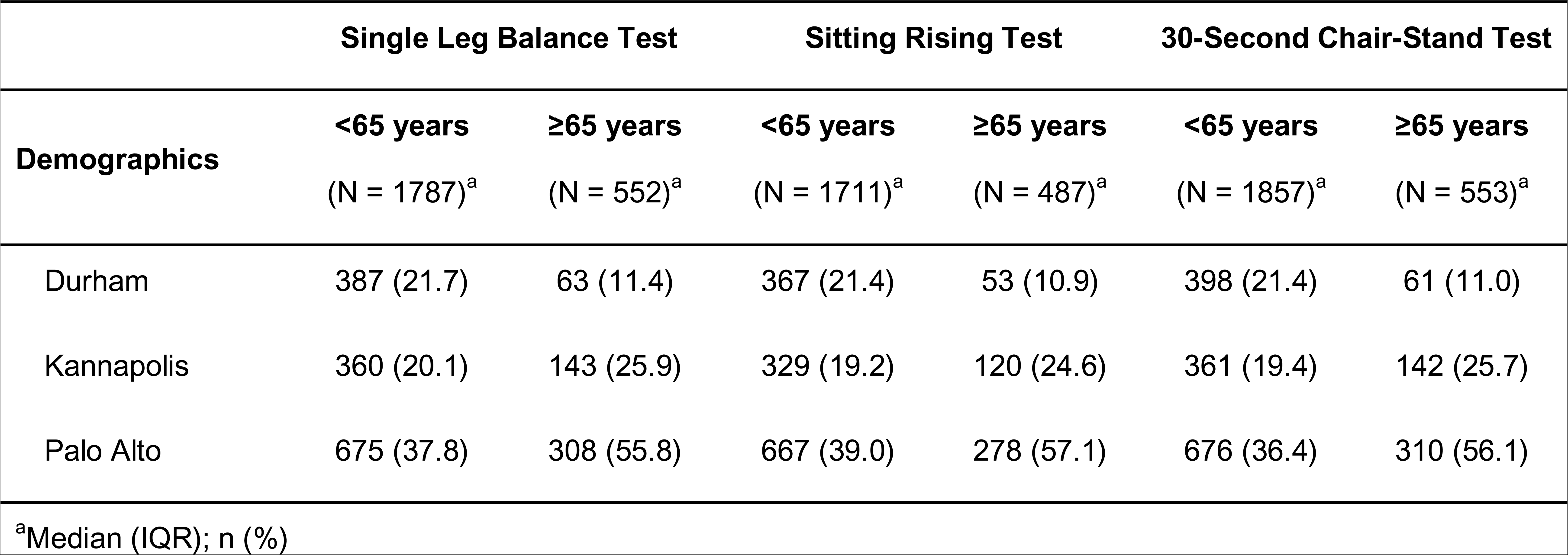
Summary of age-stratified demographic characteristics by lower quarter physical performance test.

|  | Single Leg Balance Test |  | Sitting Rising Test |  | 30-Second Chair-Stand Test |  |
| --- | --- | --- | --- | --- | --- | --- |
| Demographics | <65 years<br>(N = 1787)<sup>a</sup> | ≥65 years<br>(N = 552)<sup>a</sup> | <65 years<br>(N = 1711)<sup>a</sup> | ≥65 years<br>(N = 487)<sup>a</sup> | <65 years<br>(N = 1857)<sup>a</sup> | ≥65 years<br>(N = 553)<sup>a</sup> |
| Age, years | 43.1 (32.0,<br>54.3) | 72.3 (69.1,<br>76.4) | 42.5 (31.8,<br>53.8) | 72.2 (69.1,<br>76.3) | 42.8 (31.9,<br>54.2) | 72.3 (69.0,<br>76.4) |
| Female | 1,014 (56.7) | 282 (51.1) | 963 (56.3) | 244 (50.1) | 1,033 (55.6) | 281 (50.8) |
| Hispanic ethnicity | 256 (14.3) | 15 (2.7) | 249 (14.6) | 12 (2.5) | 264 (14.2) | 15 (2.7) |
| Race |  |  |  |  |  |  |
| White | 1,016 (56.9) | 473 (85.7) | 979 (57.2) | 415 (85.2) | 1,067 (57.5) | 474 (85.7) |
| Black or African<br>American | 329 (18.4) | 42 (7.6) | 303 (17.7) | 38 (7.8) | 332 (17.9) | 41 (7.4) |
| Asian | 214 (12.0) | 27 (4.9) | 209 (12.2) | 26 (5.3) | 224 (12.1) | 28 (5.1) |
| Native Hawaiian<br>or Other Pacific<br>Islander | 23 (1.3) | 2 (0.4) | 22 (1.3) | 2 (0.4) | 25 (1.3) | 2 (0.4) |
| American Indian<br>or Alaska Native | 24 (1.3) | 3 (0.5) | 23 (1.3) | 3 (0.6) | 24 (1.3) | 3 (0.5) |
| Other | 181 (10.1) | 5 (0.9) | 175 (10.2) | 3 (0.6) | 184 (9.9) | 5 (0.9) |
| Site |  |  |  |  |  |  |
| Los Angeles | 365 (20.4) | 38 (6.9) | 348 (20.3) | 36 (7.4) | 422 (22.7) | 40 (7.2) |
| Demographics | <65 years | ≥65 years | <65 years | ≥65 years | <65 years | ≥65 years |
|  | (N = 1787)<sup>a</sup> | (N = 552)<sup>a</sup> | (N = 1711)<sup>a</sup> | (N = 487)<sup>a</sup> | (N = 1857)<sup>a</sup> | (N = 553)<sup>a</sup> |
| Durham | 387 (21.7) | 63 (11.4) | 367 (21.4) | 53 (10.9) | 398 (21.4) | 61 (11.0) |
| Kannapolis | 360 (20.1) | 143 (25.9) | 329 (19.2) | 120 (24.6) | 361 (19.4) | 142 (25.7) |
| Palo Alto | 675 (37.8) | 308 (55.8) | 667 (39.0) | 278 (57.1) | 676 (36.4) | 310 (56.1) |

<sup>a</sup>Median (IQR); n (%)

### Main results

To focus results and avoid reporting on variables with extremely small coefficients in the final models, we present the top 20 candidate characteristics from sENET results. Models for single-legged balance and sitting-rising test scores were optimized to LASSO regressions (□=1), while models for 30CST were optimized to ENET regression (□=0.5).

#### Single-legged balance test

We present age-stratified sENET results for the SLBT in **Figure 2a** and **Figure 2b**. Across both age groups, sociodemographic factors positively associated with SLBT performance included identifying as Asian race or being married. In participants aged <65, identifying as Black or African American or having highest educational attainment being high school or less was also associated with lower odds of achieving the 60-second SLBT ceiling. Among those >65 years-old, continuous age remained an important factor negatively associated with SLBT performance.

**Figure 2.**
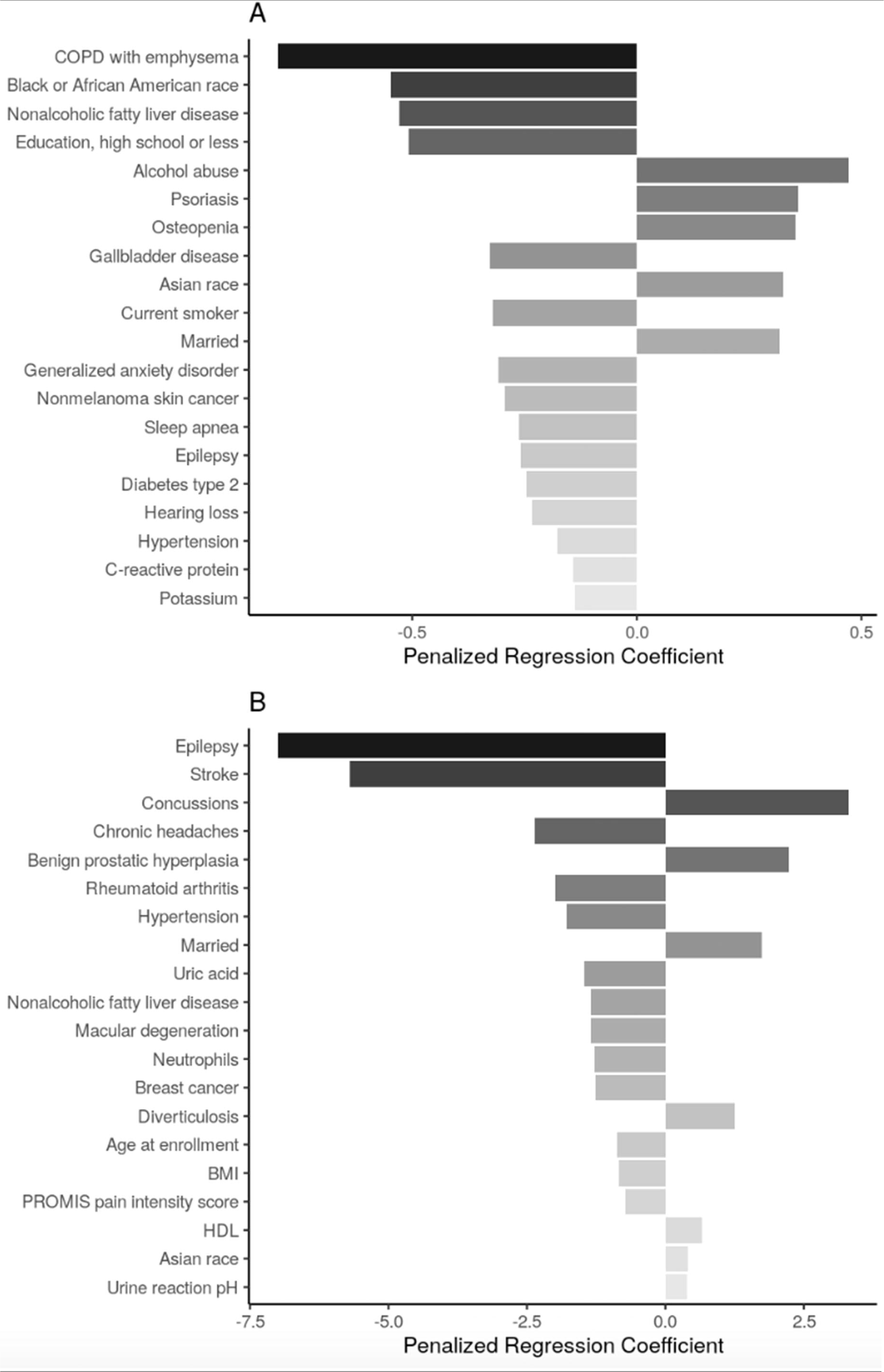
**Top 20 regression coefficients of features selected from sENET regression model for single legged balance test.** Panel A: participants <65 years of age (□ = 1, λ = 0.0025); Panel B: participants ≥65 years of age (□ = 1, λ = 0.334). Abbreviations: PTSD = posttraumatic stress disorder; BMI = body mass index, HDL = high density lipoprotein.

Health conditions and behaviors that were associated with worse SLBT performance across both age groups were non-alcoholic fatty liver disease (NAFLD), hypertension, and epilepsy. In adults <65 years-old, chronic obstructive pulmonary disease with emphysema (COPD), gallbladder disease, being a current smoker, generalized anxiety disorder, and nonmelanoma skin cancer, sleep apnea, and type II diabetes were among the factors associated with lower odds of achieving the 60-second SLBT ceiling while alcohol abuse, psoriasis, and osteopenia were among those associated with increased odds of high SLBT performance. Among adults aged ≥65 years, stroke, chronic headaches, rheumatoid arthritis, macular degeneration, and breast cancer were all associated with lower SLBT times while concussions, benign prostatic hyperplasia, and diverticulosis were associated with higher SLBT times.

Biomarkers associated with SLBT performance included C-reactive protein (CRP; associated with decreased odds of achieving 60 seconds) in adults <65 years-old. In adults aged ≥65 years, body mass index (BMI), uric acid, and neutrophil levels were negatively associated with SLBT time, while high-density lipoprotein (HDL) and urine pH levels were positively associated with SLBT time.

#### Sitting-rising test

We present age-stratified sENET results for the SRT in **Figure 3a** and **Figure 3b**. Sociodemographic factors negatively associated with SRT scores across both age subgroups were female sex and age, treated as a continuous measure beyond the stratification. Additional sociodemographic factors associated with lower SRT scores among participants aged <65 were annual income <$25,000 and identifying as Black or African American race. The only additional sociodemographic factor in the SRT model for participants aged ≥65 years was hispanic ethnicity, which was also associated with a lower SRT score.

**Figure 3.**
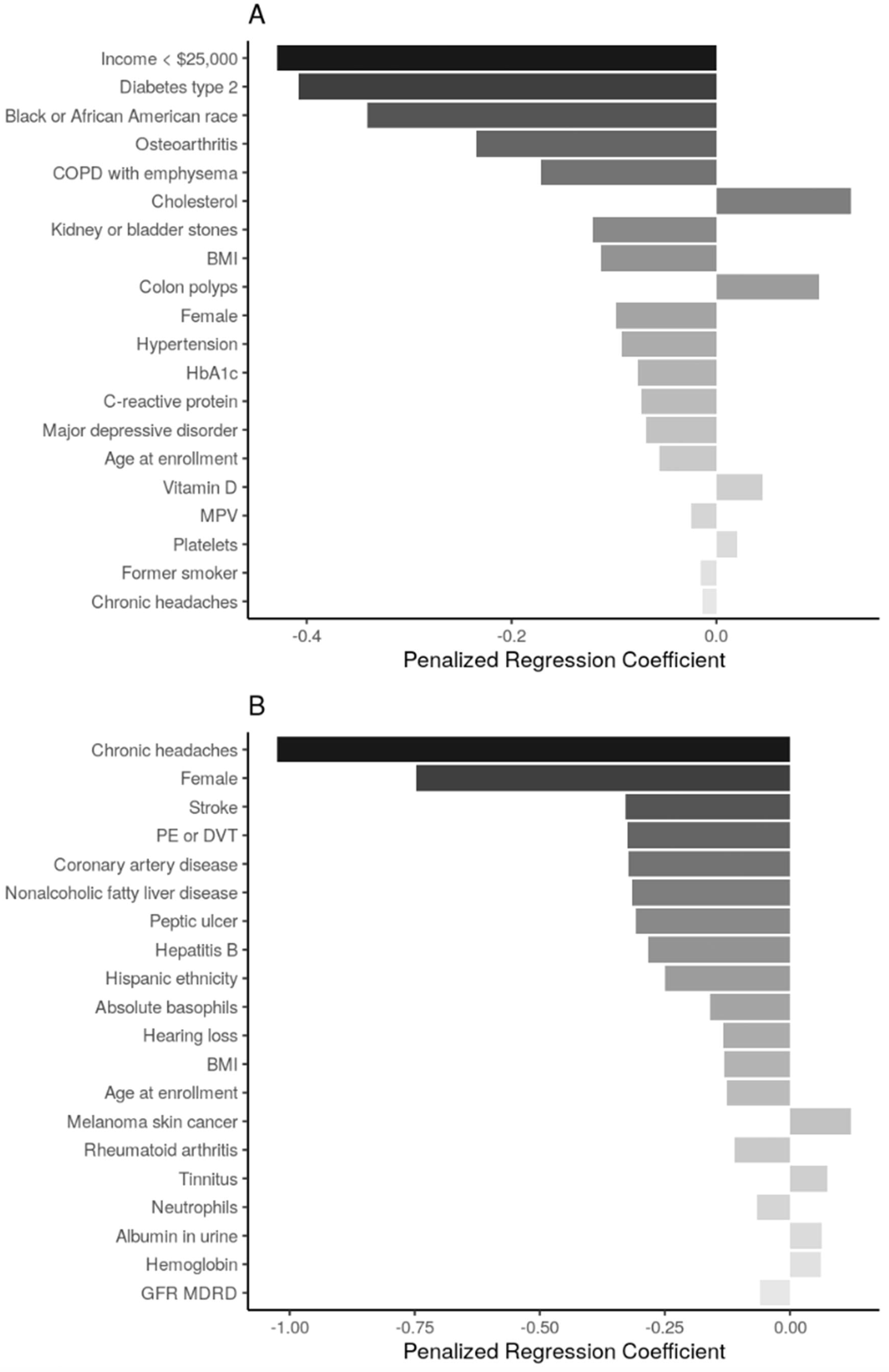
**Top 20 regression coefficients of features selected from sENET regression model for sitting rising score.** Panel A: participants <65 years of age (□ = 1, λ = 0.023); Panel B: participants ≥65 years of age (□ = 1, λ = 0.038). Abbreviations: BMI = body mass index; HbA1c = hemoglobin A1c; MPV = ; PE = pulmonary embolism; DVT = deep venous thrombosis; GFR MDRD = estimated glomerular filtration rate, Modification of Diet in Renal Disease Study Equation.

Health conditions and behaviors that were associated with worse SRT scores across both age groups were chronic headaches. Among participants aged <65 years, type 2 diabetes mellitus, osteoarthritis, COPD, hypertension, and major depressive disorder were associated with lower SRT score; conversely, colon polypswere associated with higher SRT score. Among participants aged ≥65 years, many health conditions including stroke, pulmonary embolism/deep venous thrombosis, coronary artery disease, NAFLD, peptic ulcer, and Hepatitis B were associated with lower SRT score while melanoma skin cancer and tinnitus were associated with higher SRT score.

BMI was negatively associated with SRT score in both age groups. Additional biomarkers negatively associated with SRT scores in participants <65 included HbA1c, CRP, and mean platelet volume, while cholesterol, Vitamin D, and platelet count had positive associations with SRT scores in this age group. Among participants ≥65 years, increased basophil count, neutrophil count, and glomerular filtration rate were associated with lower SRT scores, while increased urine albumin and hemoglobin were associated with higher SRT scores.

#### 30-second chair-stand test

We present age-stratified sENET results for the 30CST in **Figure 4a** and **Figure 4b**. The only sociodemographic factor shared across both age subgroups in the models for the 30CST was being Black or African American (associated with lower stand count). Among adults aged <65 years, other sociodemographic characteristics in the model were Asian race (associated with higher stand count) and being unemployed (associated with lower stand count). Among adults aged ≥65 years, additional sociodemographic factors in the 30CST model were highest education attainment being high school or less, which was associated with lower stand count, while being married was associated with higher stand count.

**Figure 4.**
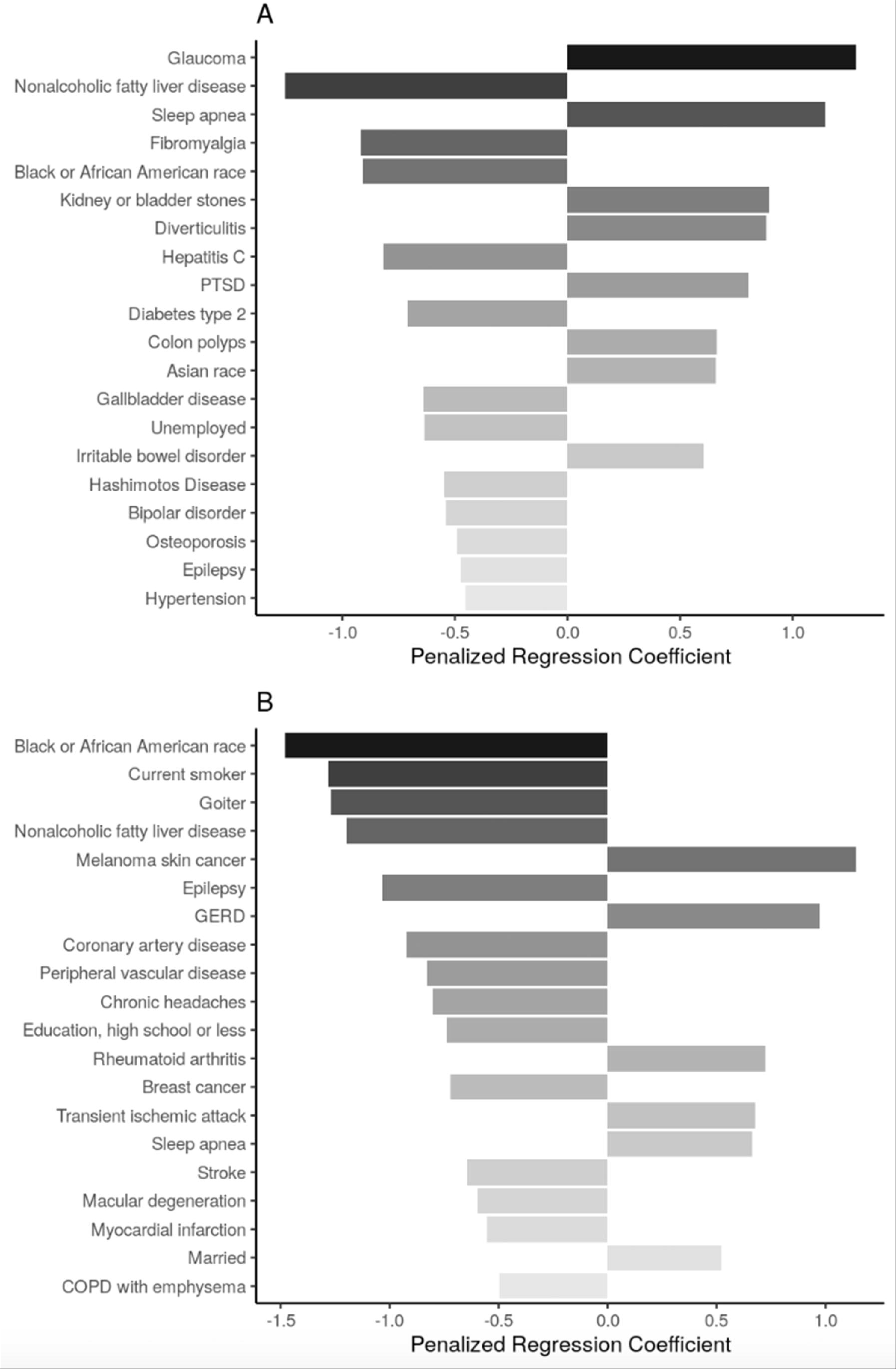
**Top 20 regression coefficients of features selected from sENET regression model for 30-second chair stand test.** Panel A: participants <65 years of age (□ = 0.5, λ = 0.0238); Panel B: participants ≥65 years of age (□ = 0.5, λ = 0.0752). Abbreviations: PTSD = posttraumatic stress disorder; GERD = gastroesophageal reflux disease; COPD = chronic obstructive pulmonary disease.

Health conditions and behaviors that were associated with lower 30CST count across both age groups were NALFD and epilepsy. Sleep apnea was also present in the models for both age subgroups, but was associated with improved 30CST performance. Among participants aged <65 years, glaucoma, kidney or bladder stones, diverticulitis, PTSD, colon polyps, and IBD were also associated with a higher stand count while fibromyalgia, hepatitis C, type 2 diabetes mellitus, gallbladder disease, and Hashimoto’s disease were among the conditions associated with lower stand count. Among participants aged ≥65 years, melanoma skin cancer, GERD, rheumatoid arthritis, and transient ischemic attack were associated with higher stand count while a variety of conditions, including being a current smoker, goiter, epilepsy, coronary artery disease, peripheral vascular disease,breast cancer, and stroke were associated with lower stand count.

Neither of the models for 30CST identified any biomarkers as one of the top 20 features selected.

### Sensitivity analysis results

We present results from sensitivity analyses in the **Supplemental Material**.

## DISCUSSION

This study identified novel associations between candidate variables and three physical performance tests using a deeply phenotyped cohort. Salient factors across performance tests and age-stratification included variables from sociodemographic characteristics, health conditions/behaviors, and biomarker values. Of note, there were a number of characteristics that were selected as top-five features across multiple performance tests and age groups. Among participants aged <65 years, being Black or African American was selected in the top five candidate characteristics for all performance tests, while among participants aged ≥65, NAFLD was consistently a top-selected feature.

Age-based reference values for SLBT, SRT and 30CST have been previously reported among community-dwelling adults (primarily those aged ≥60 years) and in disease-based populations. In general, physical function measures were slightly higher in the PBHS cohort. More specifically, the SLBT time was higher across both age groups compared to previously published data.^20,30^ For the 30CST, the mean stand count among adults aged ≥65 was only slightly higher than previously reported reference data.^30^ While direct comparison among adults aged <65 year is difficult due to availability of data, the distributions of the 30CST results were relatively similar across age-stratified groups. While age-based median values have been reported for the SRT, data availability limits direct comparison with our results.^23^

### Commonalities across sociodemographic characteristics

Several sociodemographic characteristics were associated with physical performance, for example: race and ethnicity, income level, and educational attainment. This appears to be consistent with previous research reporting that higher educational attainment is associated with a more physically active lifestyle.^31–33^ If these characteristics are causally associated with the outcomes, then general physical activity would impact the physical performance measures of interest in our study. Substantial disparities in physical activity among adolescents and young adults by race and income have also been previously reported.^34^ The relationship between race and physical function in our study may be explained by the impact of racial and ethnic bias on health care delivery in the US,^35^ the breadth of racial disparities in health outcomes,^35,36^ and how such disparities may impact physical ability captured via physical performance tests. Our results in the context of prior research underline the complex relationship between physical performance ability with sociodemographics and the social determinants of health. Additional research is warranted to identify whether targeted exercise and physical activity interventions impact the associations observed between physical performance and sociodemographic factors in our study.

### Commonalities across health conditions and behaviors

Our results confirm previous findings associating reduced physical function and with disease and health behaviors. Some associations highlight the potential to identify subclinical disease in patients at risk. For example, both SLBT and SRT performance were associated with hypertension among adults aged <65 and a history of several cardiovascular conditions and events (e.g., stroke, coronary artery disease) among adults aged ≥65; SLBT and SRT performance in early and/or middle-adulthood may have value in predicting future cardiovascular disease conditions or events later in life.

NAFLD was notably associated with worse performance on the SLBT, SRT, and 30CST and was in the top 5 factors overall associated with each test across both age groups for the SLBT and 30CST. It was also among the top 5 factors associated with the SRT for those aged ≥65 and in the top 10 factors for those aged <65. NAFLD is prevalent in a little more than one-third of the general North and South American population^37^ and is over two-times more prevalent among adults who are overweight (75%) or obese (76%).^38^ Validation of the value the SLBT, SRT, and 30CST have in low-cost screening to identify individuals who need further diagnostic testing for NAFLD is needed in future research.

Among those aged ≥65, each of the physical performance measures was negatively associated with experiencing chronic headaches. Although previous research indicates chronic headaches tend to have negative effects on physical activity levels,^39^ it is unclear why this consistent relationship was limited to those ≥65 years-old. Future research is needed to validate and further illuminate the nuance of this association.

### Commonalities across biomarkers

Biomarkers associated with each of the three physical performance tests had limited commonalities across tests and age-groups. While only associated with two physical performance tests in the younger population (SLBT and SRT), CRP was shown to be negatively associated with each test. CRP is commonly used as a marker of systemic inflammation,^40^ with higher levels indicating acute inflammatory responses or infectious disease - both of which can impact physical performance. CRP may also indicate chronic inflammation associated with other disease states, such as atherosclerotic cardiovascular disease.^41^

We also found a negative association between BMI and SLBT scores in the older population, as well as SRT scores in both age groups. Prior research has similarly reported on an inverse relationship between body composition and physical function measures.^42–44^

### Limitations

First, the purpose of this study and analysis approach we used are exploratory and cross-sectional in nature, with the goal of identifying the most salient factors associated with each physical function measure out of a wide list of candidate variables. Associations identified in our results (both their magnitude and direction) should not be interpreted as causal effects.^45^ Follow-up studies are needed to determine and validate the individual diagnostic or prognostic predictive value SLBT, SRT, and 30CST as they relate to the novel associations identified in our study. Any desire to estimate related causal effects will require an alternative methodological approach.^46–48^ Second, while the PBHS participants are overall representative of U.S. adult age, sex, race, and ethnicity^18^ and were recruited and enrolled at multiple sites, these sites were limited to two states (California and North Carolina) which may limit our ability to generalize these results to adults in other regions of the U.S..

In this study, we identified several novel salient factors with the SLBT, SRT, and 30CST when stratifying by age (<65 and ≥65 years) - many of them common across the three tests. Our results support the need for further investigation of the potential for these physical performance tests as affordable, noninvasive biomarkers of prevalent conditions and their predictive utility for incident health conditions in later life (e.g., cardiovascular disease and/or events) in community-dwelling adults. Further research would also be warranted to estimate the direction and magnitude of any causal relationships between these physical function tests and novel factors identified where biologically plausible.

## Data Availability

The deidentified PBHS data corresponding to this study are available upon request for the purpose of examining its reproducibility. Interested investigators should direct requests to. Requests are subject to approval by PBHS governance.

## ACKNOWLEDGEMENTS

*Project Baseline Health Study Team:* American Society of Clinical Oncology, Alexandria, VA, USA: Richard L. Schilsky. Duke University, School of Medicine, Durham, NC, USA: Jennifer Allen, MaryAnn Anderson, Kevin Anstrom, Lucus Araujo, Kristine Arges, Kaveh Ardalan, Bridget Baldwin, Suresh Balu, Mustafa R. Bashir, Manju Bhapkar, Robert Bigelow, Tanya Black, Rosalia Blanco, Gerald Bloomfield, Durga Borkar, Leah Bouk, Ebony Boulware, Nikki Brugnoni, Erin Campbell, Paul Campbell, Larry Carin, Tammy Jo Cassella, Tina Cates, Ranee Chatterjee Montgomery, Victoria Christian, John Choong, Michael Cohen-Wolkowiez, Elizabeth Cook, Scott Cousins, Ashley Crawford, Nisha Datta, Melissa Daubert, James Davis, Jillian Dirkes, Isabelle Doan, Marie Dockery, P. Murali Doraiswamy, Pamela S. Douglas, Shelly Duckworth, Ashley Dunham, Gary Dunn, Ryan Ebersohl, Julie Eckstrand, Vivienne Fang, April Flora, Emily Ford, Lucia Foster, Elizabeth Fraulo, John French, Geoffrey S. Ginsburg, Cindy Green, Latoya Greene, Jeffrey Guptill, Donna Hamel, Jennifer Hamill, Chris Harrington, Rob Harrison, Lauren Hedges, Brooke Heidenfelder, Adrian F. Hernandez, Cindy Heydary, Tim Hicks, Lina Hight, Deborah Hopkins, Erich S. Huang, Grace Huh, Jillian Hurst, Kelly Inman, Gemini Janas, Glenn Jaffee, Janace Johnson, Tiffanie Keaton, Michel Khouri, Daniel King, Jennifer Korzekwinski, Lynne H. Koweek, Anthony Kuo, Lydia Kwee, Dawn Landis, Rachele Lipsky, Desiree Lopez, Carolyn Lowry, Kelly Marcom, Keith Marsolo, Paige McAdams, Shannon McCall, Robert McGarrah, John McGugan, Dani Mee, Sabrena Mervin-Blake, Prithu Mettu, Mathias Meyer, Justin Meyers, Calire N. Miller, Rebecca Moen, Lawrence H. Muhlbaier, Michael Murphy, Ben Neely, L. Kristin Newby, Jayne Nicoldson, Hoang Nguyen, Maggie Nguyen, Lori O’Brien, Sumru Onal, Jeremey O’Quinn, David Page, Neha J. Pagidipati, Kishan Parikh, Sarah R. Palmer, Bray Patrick-Lake, Brenda Pattison, Michael Pencina, Eric D. Peterson, Jon Piccini, Terry Poole, Tom Povsic, Alicia Provencher, Dawn Rabineau, Annette Rich, Susan Rimmer, Fides Schwartz, Angela Serafin, Nishant Shah, Svati Shah, Kelly Shields, Steven Shipes, Peter Shrader, Jon Stiber, Lynn Sutton, Geeta Swamy, Betsy Thomas, Sandra Torres, Debara Tucci, Anthony Twisdale, Susan A. Whitney, Robin Williamson, Lauren Wilverding, Charlene A. Wong, Lisa Wruck. Ellen Young Gemini Group, USA: Jane Perlmutter. Health Collaboratory and Cancer 101, New York, NY, USA: Sarah Krug. Rare Dots, Inc., USA: S. Whitney Bowman-Zatzkin. Society of Participatory Medicine, USA: Sarah Krug. Stanford University, School of Medicine, Stanford, CA, USA: Themistocles Assimes, Vikram Bajaj, Maxwell Cheong, Millie Das, Manisha Desai, Alice C. Fan, Dominik Fleischmann, Sanjiv S. Gambhir, Garry Gold, Francois Haddad, David Hong, Curtis Langlotz, Yaping J. Liao, Rong Lu, Kenneth W. Mahaffey, David Maron, Rebecca McCue, Rajan Munshi, Fatima Rodriguez, Sumana Shashidhar, George Sledge, Susie Spielman, Ryan Spitler, Sue Swope, Donna Williams, Julio C. Nunes. University of Florida, College of Medicine, Gainesville, FL, USA: Carl J Pepine. University of Missouri, Children’s Mercy Hospital, Kansas City, MO, USA: John D Lantos. University of Texas, Dell Medical School, Austin, TX, USA: Michael Pignone. University of Washington, Department of Biostatistics, Seattle, WA, USA: Patrick Heagerty. Vanderbilt University, School of Medicine, Nashville, TN, USA: Laura Beskow, Gordon Bernard. Verily Inc., South San Francisco, CA, USA: Kelley Abad, Giulia Angi, Robert M. Califf, Lawrence Deang, Joy Huynh, Manway Liu, Cherry Mao, Michael Magdaleno, William J. Marks, Jr., Jessica Mega, David Miller, Nicole Ong, Darshita Patel, Vanessa Ridaura, Scarlet Shore, Sarah Short, Michelle Tran, Veronica Vu, Celeste Wong. Harvard University, School of Medicine, Boston, MA, USA: Robert C. Green. Google Inc., Mountain View, CA, USA: John Hernandez. California Health and Longevity Institute, Westlake Village, CA, USA: Jolene Benge, Gislia Negrete, Gelsey Sierra, Terry Schaack

## Conflict of Interest

MKC and SS are employees of Verily Life Sciences. The other authors report no relevant disclosures.

## Author Contributions

*Conceptualization:* Taylor, Carroll.

*Methodology*: Taylor, Carroll, Short.

*Validation*: Carroll, Short.

*Formal analysis*: Carroll.

*Data Curation:* Carroll, Short.

*Writing - Original Draft:* Taylor, Carroll, Cauwenberghs.

*Writing - Review & Editing*: Taylor, Carroll, Short, Celestin, Gilbertson, Olivier, Haddad, Cauwenberghs.

*Visualization:* Carroll.

*Technical or material support:* Short, Celestin, Gilbertson.

*Supervision and project administration:* Taylor.

## Sponsor’s Role

The employees from the funding source contributed to the data analyses, interpretation, editing of the manuscript and are coauthors. The final decision to submit the manuscript was made by the academic authors.

## Notes

### Competing Interest Statement

The Project Baseline Health Study and this analysis are funded by Verily Life Sciences, San Francisco, CA. This analysis was funded by Verily Life Sciences (San Francisco, CA) in partnership with Stanford University and Duke University. The employees from the funding source contributed to the data analyses, interpretation, editing of the manuscript and are coauthors. The final decision to submit the manuscript was made by the academic authors.

### Funding Statement

The Project Baseline Health Study and this analysis are funded by Verily Life Sciences, San Francisco, CA.

### Author Declarations

The PBHS is a multicenter longitudinal cohort study of adults in the United States (ClinicalTrials.gov identifier NCT03154346), approved by both a central Institutional Review Board (the WCG IRB; approval tracking number 20170163, work order number 1-1506365-1) and IRBs at each of the participating institutions: Stanford University (Palo Alto, CA), Duke University (Durham, NC; Kannapolis, NC), and the California Health and Longevity Institute (CHLI; Los Angeles, CA).

