## Supplemental Materials for "Identifying Factors Associated with Lower Quarter Performance-Based Balance and Strength Tests: the Project Baseline Health Study"

### Stacked Elastic Net

Stacked objective functions were chosen to handle multiply imputed data; imputed datasets are stacked on top of each other to estimate regression coefficients. The inflated sample size is addressed by using observation weights.^1^ Elastic net (ENET) regularization methods use two regression parameters to select main effects - α and λ - while considering correlation between model predictors. For this analysis, the α (L1 regularization) parameter was optimized across values of 0.5 or 1. The λ (L2 regularization) parameter was optimized across 100 values; the maximum value (λ_max_) was chosen as the smallest λ based on a model where all coefficients were equal to 0, and the minimum was equivalent to λ_max_*10^-3^. Optimal α and λ values were selected as those with the largest L1 penalty (ɑλ) using 5-fold cross validation via the cv.saenet function in the {miselect} package.^29^ These methods standardize all variables to a normal (N ~ (0, 1)) distribution, facilitating meaningful comparison across coefficient values within each subgroup analysis. Due to limitations in computation power, we only considered alpha values equivalent to 0.5 or 1; we may have considered a broader range of parameter values for model optimization given more computational power.

### Sensitivity Analyses

Health characteristic/behavior data, including symptoms related to physical health, were collected, but omitted from the main analysis as they were less easily discernible. We performed a sensitivity analysis including these data in addition to those in the main analysis; a full list of covariates can be found in **Supplemental Table 1.** It is important to note that coefficients from the same physical performance test/age-subgroup combination are not directly comparable in terms of magnitude for two reasons: 1) the sENET regularization optimization approach (described above) is specific to the data being input into the model and 2) the standardization of coefficients that allows for comparison of magnitude of association *within* each test-and-age-group specific model occurs after sENET selects the most salient covariate associations.

### Sensitivity Analyses Results

Because sENET regularization optimization is specific to the data being input into the model, adding additional variables into the modeling to be considered for sensitivity analyses resulted in small differences in optimal λ between our primary results and our sensitivity analysis results.

**Adding self-reported symptoms**. Sensitivity analyses adding symptoms to the pool of candidate variables did not substantially change the most salient features associated with the physical function test performance. We present these results in **Figure S1**, **Figure S2**, **Figure S3**.

**TABLE**

**Supplemental Table 1.** List of candidate covariates for multivariable ENET regression (inclusion indicated by **X**). All data collected at baseline study visit or within the first 200 days of enrollment.

|  |  | **Model 1** | | **Supplemental Model 1** | |
| --- | --- | --- | --- | --- | --- |
| **Variable Name** | **Definition** | Age < 65 | Age ≥ 65 | Age < 65 | Age ≥ 65 |
| **Demographic Characteristics** | | | | | |
| Age | Age in years | **X** | **X** | **X** | **X** |
| Sex | Self-reported sex at birth | **X** | **X** | **X** | **X** |
| Race | Self-reported race | **X** | **X** | **X** | **X** |
| Ethnicity | Self-reported Hispanic ancestry (yes/no) | **X** | **X** | **X** | **X** |
| Smoking Status | Self-reported smoking status | **X** | **X** | **X** | **X** |
| Pack-Years Smoked | Self-reported cigarettes smoked per day multiplied by years of regular smoking divided by 20 | **X** | **X** | **X** | **X** |
| **SES-related Characteristics via Life Circumstances and Habits Survey** | | | | | |
| Highest Education Completed | Self-reported highest education level completed from first survey completed | **X** | **X** | **X** | **X** |
| Household Income | Self-reported household income from first survey completed | **X** | **X** | **X** | **X** |
| Marital Status | Self-reported marital status from first survey completed | **X** | **X** | **X** | **X** |
| Employment Status | Self-reported employment status from first survey completed | **X** | **X** | **X** | **X** |
| Health Insurance | Self-reported health insurance (yes/no) from first survey completed | **X** | **X** | **X** | **X** |
| **Physical Screening** | | | | | |
| BMI | Body Mass Index (kg/m^2^) | **X** | **X** | **X** | **X** |
| **Participant Reported Physical Health-Related Medical Conditions** | | | | | |
| ADHD | Self-reported attention deficit hyperactivity disorder at baseline | **X** | **X** | **X** | **X** |
| Alcohol Abuse | Self-reported alcohol abuse at baseline | **X** | **X** | **X** | **X** |
| Arrhythmia | Self-reported arrhythmia at baseline | **X** | **X** | **X** | **X** |
| Asthma | Self-reported asthma at baseline | **X** | **X** | **X** | **X** |
| Atrial fibrillation | Self-reported atrial fibrillation |  | **X** |  | **X** |
| Benign Prostatic Hyperplasia | Self-reported benign prostatic hyperplasia at baseline |  | **X** |  | **X** |
| Bipolar Disorder | Self-reported bipolar disorder at baseline | **X** |  | **X** |  |
| Breast Cancer | Self-reported breast cancer at baseline |  | **X** |  | **X** |
| Cataracts | Self-reported cataracts at baseline | **X** | **X** | **X** | **X** |
| Chronic Headaches | Self-reported chronic headaches (non-migraines) at baseline | **X** | **X** | **X** | **X** |
| Chronic Obstructive Pulmonary Disease (COPD) | Self-reported COPD (with emphysema) at baseline | **X** | **X** | **X** | **X** |
| Colon Polyps | Self-reported colon polyps at baseline | **X** | **X** | **X** | **X** |
| Concussion or Loss of Consciousness | Self-reported concussion or loss of consciousness at baseline | **X** | **X** | **X** | **X** |
| Coronary Artery Disease | Self-reported coronary artery disease (including angina) at baseline | **X** | **X** | **X** | **X** |
| Diabetes type 1 | Self-reported Diabetes type 1 | **X** |  | **X** |  |
| Diabetes type 2 | Self-reported Diabetes type 2 | **X** | **X** | **X** | **X** |
| Diverticulitis | Self-reported diverticulitis at baseline | **X** | **X** | **X** | **X** |
| Diverticulosis | Self-reported diverticulosis at baseline | **X** | **X** | **X** | **X** |
| Drug Abuse | Self-reported drug abuse (including prescription medications) at baseline | **X** |  | **X** |  |
| Epilepsy | Self-reported epilepsy | **X** | **X** | **X** | **X** |
| Fibromyalgia | Self-reported fibromyalgia at baseline | **X** |  | **X** |  |
| GAD | Self-reported generalized anxiety disorder at baseline | **X** | **X** | **X** | **X** |
| Gallbladder Disease | Self-reported gallbladder disease at baseline | **X** | **X** | **X** | **X** |
| GERD | Self-reported gastroesophageal reflux disease at baseline | **X** | **X** | **X** | **X** |
| Glaucoma | Self-reported glaucoma at baseline | **X** | **X** | **X** | **X** |
| Goiter | Self-reported goiter |  | **X** |  | **X** |
| Gout | Self-reported gout at baseline | **X** | **X** | **X** | **X** |
| Hashimoto’s Disease | Self-reported Hashimoto’s disease | **X** | **X** | **X** | **X** |
| Hemorrhoids | Self-reported hemorrhoids at baseline | **X** | **X** | **X** | **X** |
| Hepatitis B | Self-reported Hepatitis B |  | **X** |  | **X** |
| Hepatitis C | Self-reported Hepatitis C | **X** |  | **X** |  |
| Hypercholesterolemia | Self-reported hypercholesterolemia at baseline | **X** | **X** | **X** | **X** |
| Hyperlipidemia | Self-reported hyperlipidemia at baseline | **X** | **X** | **X** | **X** |
| Hypertension | Self-reported hypertension at baseline | **X** | **X** | **X** | **X** |
| Hypothyroidism | Self-reported hypothyroidism at baseline | **X** | **X** | **X** | **X** |
| Insomnia | Self-reported insomnia at baseline | **X** | **X** | **X** | **X** |
| Irritable Bowel Disorder | Self-reported irritable bowel disorder at baseline | **X** | **X** | **X** | **X** |
| Kidney or Bladder Stones | Self-reported kidney or bladder stones at baseline | **X** | **X** | **X** | **X** |
| Macular degeneration | Self-reported macular degeneration |  | **X** |  | **X** |
| MDD | Self-reported major depressive disorder at baseline | **X** | **X** | **X** | **X** |
| Melanoma Skin Cancer | Self-reported melanoma skin cancer at baseline |  | **X** |  | **X** |
| Migraines | Self-reported migraine headaches at baseline | **X** | **X** | **X** | **X** |
| Myocardial Infarction | Self-reported myocardial infarction at baseline |  | **X** |  | **X** |
| Non-alcoholic fatty liver disease | Self-reported non-alcoholic fatty liver disease | **X** | **X** | **X** | **X** |
| Non-melanoma skin cancer | Self-reported non-melanoma skin cancer at baseline | **X** | **X** | **X** | **X** |
| Osteoarthritis | Self-reported osteoarthritis at baseline | **X** | **X** | **X** | **X** |
| Osteopenia | Self-reported osteopenia at baseline | **X** | **X** | **X** | **X** |
| Osteoporosis | Self-reported osteoporosis at baseline | **X** | **X** | **X** | **X** |
| Peptic Ulcers | Self-reported peptic ulcers at baseline | **X** | **X** | **X** | **X** |
| Peripheral vascular disease | Self-reported peripheral vascular disease |  | **X** |  | **X** |
| Pneumonia | Self-reported pneumonia | **X** | **X** | **X** | **X** |
| Prostate cancer | Self-reported prostate cancer |  | **X** |  | **X** |
| Psoriasis | Self-reported psoriasis at baseline | **X** | **X** | **X** | **X** |
| PTSD | Self-reported post-traumatic stress disorder at baseline | **X** |  | **X** |  |
| Pulmonary Embolism (PE) or Deep Vein Thrombosis (DVT) | Self-reported PE or DVT at baseline |  | **X** |  | **X** |
| Rheumatoid Arthritis | Self-reported rheumatoid arthritis at baseline | **X** | **X** | **X** | **X** |
| Severe Hearing Loss | Self-reported severe hearing loss at baseline | **X** | **X** | **X** | **X** |
| Sleep Apnea | Self-reported sleep apnea at baseline | **X** | **X** | **X** | **X** |
| Stroke | Self-reported stroke |  | **X** |  | **X** |
| Tinnitus | Self-reported tinnitus at baseline | **X** | **X** | **X** | **X** |
| Transient ischemic attack | Self-reported transient ischemic attack |  | **X** |  | **X** |
| **Physical Health-Related Symptoms** | | | | | |
| Appetite Changes | Self-reported appetite changes at baseline |  |  | **X** | **X** |
| Backache | Self-reported backache at baseline |  |  | **X** | **X** |
| Bloating | Self-reported bloating at baseline |  |  | **X** | **X** |
| Body Image Concerns | Self-reported body image concerns at baseline |  |  | **X** | **X** |
| Constipation | Self-reported constipation at baseline |  |  | **X** | **X** |
| Cough | Self-reported cough at baseline |  |  | **X** | **X** |
| Coughing up Sputum | Self-reported coughing up sputum at baseline |  |  | **X** | **X** |
| Cramping | Self-reported cramping at baseline |  |  | **X** | **X** |
| Diarrhea | Self-reported diarrhea at baseline |  |  | **X** | **X** |
| Difficulty Concentrating | Self-reported difficulty concentrating at baseline |  |  | **X** | **X** |
| Discharge | Self-reported discharge at baseline |  |  | **X** | **X** |
| Dry Mouth | Self-reported dry mouth at baseline |  |  | **X** | **X** |
| Dryness | Self-reported dryness at baseline |  |  | **X** | **X** |
| Ear Ringing | Self-reported ear ringing at baseline |  |  | **X** | **X** |
| Easy Bruising or Bleeding | Self-reported easy bruising or bleeding at baseline |  |  | **X** | **X** |
| Excessive Belching or Passing of Gas | Self-reported excessive belching or Passing of gas at baseline |  |  | **X** | **X** |
| Fatigue | Self-reported fatigue at baseline |  |  | **X** | **X** |
| Floaters | Self-reported floaters at baseline |  |  | **X** | **X** |
| Frequency of Urination | Self-reported frequency of urination at baseline |  |  | **X** | **X** |
| Hay Fever | Self-reported hay fever at baseline |  |  | **X** | **X** |
| Headache | Self-reported headache at baseline |  |  | **X** | **X** |
| Heartburn | Self-reported heartburn at baseline |  |  | **X** | **X** |
| Heat or Cold Intolerance | Self-reported heat or cold intolerance at baseline |  |  | **X** | **X** |
| Hemorrhoids | Self-reported hemorrhoids at baseline |  |  | **X** | **X** |
| Itching | Self-reported itching at baseline |  |  | **X** | **X** |
| Joint Pain Swelling | Self-reported joint pain swelling at baseline |  |  | **X** | **X** |
| Lack of Energy | Self-reported lack of energy at baseline |  |  | **X** | **X** |
| Leg Cramps | Self-reported leg cramps at baseline |  |  | **X** | **X** |
| Lightheadedness | Self-reported lightheadedness at baseline |  |  | **X** | **X** |
| Memory Change | Self-reported memory change at baseline |  |  | **X** | **X** |
| Mood Change | Self-reported mood change at baseline |  |  | **X** | **X** |
| Muscle or Joint Pain | Self-reported muscle or joint pain at baseline |  |  | **X** | **X** |
| Nasal Stuffiness | Self-reported nasal stuffiness at baseline |  |  | **X** | **X** |
| Neck or Low Back Pain | Self-reported neck or low back pain at baseline |  |  | **X** | **X** |
| Nervousness | Self-reported nervousness at baseline |  |  | **X** | **X** |
| Night Sweats | Self-reported night sweats at baseline |  |  | **X** | **X** |
| Numbness or Loss of Sensation | Self-reported numbness or loss of sensation at baseline |  |  | **X** | **X** |
| Pain or Stiffness in the Neck | Self-reported pain or stiffness in neck at baseline |  |  | **X** | **X** |
| Runny Nose | Self-reported runny nose at baseline |  |  | **X** | **X** |
| Shortness of Breath | Self-reported shortness of breath at baseline |  |  | **X** | **X** |
| Shortness of Breath with Exercise | Self-reported shortness of breath with exercise at baseline |  |  | **X** | **X** |
| Sinus Pain | Self-reported sinus pain at baseline |  |  | **X** | **X** |
| Sleeping Pattern Changes | Self-reported sleeping pattern changes at baseline |  |  | **X** | **X** |
| Stiffness | Self-reported stiffness at baseline |  |  | **X** | **X** |
| Swelling in Calves or Feet | Self-reported swelling in calves or feet at baseline |  |  | **X** | **X** |
| Tension | Self-reported tension at baseline |  |  | **X** | **X** |
| Tingling or Numbness in Extremities | Self-reported tingling or numbness in extremities at baseline |  |  | **X** | **X** |
| Tingling or Pins and Needles | Self-reported tingling or pins and needles at baseline |  |  | **X** | **X** |
| Urgency | Self-reported urgency at baseline |  |  | **X** | **X** |
| Urination at Night | Self-reported urination at night at baseline |  |  | **X** | **X** |
| Any food allergies | Self-reported food allergies (any vs. none, ignoring additional details about which allergen) |  |  | **X** | **X** |
| Any seasonal allergies | Self-reported seasonal allergies (any vs. none, ignoring additional details about which allergen) |  |  | **X** | **X** |
| Any non-seasonal allergies | Self-reported non-seasonal allergies (any vs. none, ignoring additional details about which allergen) |  |  | **X** | **X** |
| Any medication allergies | Self-reported medication allergies (any vs. none, ignoring additional details about which allergen) |  |  | **X** | **X** |
| **Blood Draw - Standard Laboratory Data** | | | | | |
| Hemoglobin | Hemoglobin (g/dl) at baseline | **X** | **X** | **X** | **X** |
| Serum Creatinine | Serum Creatinine (mg/dl) at baseline | **X** | **X** | **X** | **X** |
| HDL | High density lipoprotein (mg/dl) at baseline | **X** | **X** | **X** | **X** |
| LDL | Low density lipoprotein (mg/dl) at baseline | **X** | **X** | **X** | **X** |
| Triglycerides | Triglycerides (mg/dl) at baseline | **X** | **X** | **X** | **X** |
| HbA1c | Hemoglobin A1c (%) at baseline | **X** | **X** | **X** | **X** |
| Neutrophils | Neutrophils (k/mcL) at baseline | **X** | **X** | **X** | **X** |
| Lymphocytes | Lymphocytes (k/mcL) at baseline | **X** | **X** | **X** | **X** |
| ALT | Alanine aminotransferase (U/L) at baseline | **X** | **X** | **X** | **X** |
| AST | Aspartate aminotransferase (U/L) at baseline | **X** | **X** | **X** | **X** |
| Vitamin D | Vitamin D (ng/ml) at baseline | **X** | **X** | **X** | **X** |
| CRP | C-reactive protein (mg/l) at baseline | **X** | **X** | **X** | **X** |
| Blood Glucose | Blood glucose (mg/dl) at baseline | **X** | **X** | **X** | **X** |
| Neutrophil Segments | Neutrophil segments (% WBC) at baseline | **X** | **X** | **X** | **X** |
| Magnesium | Magnesium (MEQ/L) at baseline | **X** | **X** | **X** | **X** |
| Hematocrit | Hematocrit (% RBC to whole blood volume) at baseline | **X** | **X** | **X** | **X** |
| MCV | Mean corpuscular volume (fL) at baseline | **X** | **X** | **X** | **X** |
| MCH | Mean corpuscular hemoglobin (pg) at baseline | **X** | **X** | **X** | **X** |
| MPV | Mean platelet volume (fL) at baseline | **X** | **X** | **X** | **X** |
| Platelet Count | Platelet count (cumm) at baseline | **X** | **X** | **X** | **X** |
| RBC Count | Red blood cell count (millions/mcL) at baseline | **X** | **X** | **X** | **X** |
| WBC Count | White blood cell count (thousands/mcL) at baseline | **X** | **X** | **X** | **X** |
| Calcium | Calcium (mg/dL) at baseline | **X** | **X** | **X** | **X** |
| Cholesterol | Total cholesterol (mg/dL) at baseline | **X** | **X** | **X** | **X** |
| Chloride | Chloride (MEQ/L) at baseline | **X** | **X** | **X** | **X** |
| Potassium | Potassium (MEQ/L) at baseline | **X** | **X** | **X** | **X** |
| Sodium | Sodium (MEQ/L) at baseline | **X** | **X** | **X** | **X** |
| Protein (Serum) | Protein in serum (g/dL) at baseline | **X** | **X** | **X** | **X** |
| Uric Acid | Uric acid (mg/dL) at baseline | **X** | **X** | **X** | **X** |
| Absolute Monocytes | Absolute Monocytes (k/mcL) at baseline | **X** | **X** | **X** | **X** |
| Absolute Eosinophils | Absolute Eosinophils (k/mcL) at baseline | **X** | **X** | **X** | **X** |
| Absolute Basophils | Absolute Basophils (k/mcL) at baseline | **X** | **X** | **X** | **X** |
| Creatinine (Urine) | Creatinine in urine (mg/dL) at baseline | **X** | **X** | **X** | **X** |
| GFR MDRD | Glomerular filtration rate (mL/min/1.73 m^2^) based on Modification of Diet in Renal Disease Study equation at baseline | **X** | **X** | **X** | **X** |
| Absolute Reticulocytes | Absolute reticulocytes (billions/L) at baseline | **X** | **X** | **X** | **X** |
| TSH | Thyroid stimulating hormone (mIU/L) at baseline | **X** | **X** | **X** | **X** |
| Urine Reaction pH | Urine reaction pH at baseline | **X** | **X** | **X** | **X** |
| Urine Specific Gravity | Urine specific gravity at baseline | **X** | **X** | **X** | **X** |
| Albumin | Albumin (g/L) at baseline | **X** | **X** | **X** | **X** |
| **Patient Reported Outcome Assessments** | | | | | |
| Sheehan Disability Scale Score | Sheen Disability Scale total score (range 0, 30) at baseline | **X** | **X** | **X** | **X** |
| WHODAS 2.0 Score | WHODAS 2.0 total score (range 0, 48) at baseline | **X** | **X** | **X** | **X** |
| PHQ-9 Score | Patient Health Questionnaire-9 total score (range 0, 27) at baseline | **X** | **X** | **X** | **X** |
| GAD-7 Score | Generalized Anxiety Disorder-7 total score (range 0, 21) at baseline | **X** | **X** | **X** | **X** |
| BRFSS ACE Score | Behavioral Risk Factor Surveillance System Adverse Childhood Experience Module total problem count (range 0, 11) at baseline | **X** | **X** | **X** | **X** |
| PANAS Mood Positive Affect Score | PANAS Mood Positive Affect total score (range 10, 50) from first survey completed | **X** | **X** | **X** | **X** |
| PANAS Mood Negative Affect Score | PANAS Mood Positive Affect total score (range 10, 50) from first survey completed | **X** | **X** | **X** | **X** |
| Satisfaction with Life Score | Satisfaction with Life total score (range 5, 35) from first survey completed | **X** | **X** | **X** | **X** |
| Subjective Happiness Score | Subjective Happiness total score (range 4, 28) from first survey completed | **X** | **X** | **X** | **X** |
| AUDIT-C Score | Alcohol Use Disorders Identification Test-Concise total score (range 0, 12) from first survey completed | **X** | **X** | **X** | **X** |
| PROMIS Pain Intensity Score | PROMIS Pain Intensity total score (range 3, 15) from first survey completed | **X** | **X** | **X** | **X** |
| PROMIS Pain Interference Score | PROMIS Pain Interference total score (range 6, 30) from first survey completed | **X** | **X** | **X** | **X** |
| Perceived Social Support Score | Perceived Social Support total score (range 12, 84) at baseline from first survey completed | **X** | **X** | **X** | **X** |

**FIGURES**

**Figure S1.** Top 20 regression coefficients of features selected from sENET regression model for single legged balance test, with the addition of features related to symptoms and allergies. Panel A: participants <65 years of age (ɑ = 1, λ = 0.0041); Panel B: participants ≥65 years of age (ɑ = 1, λ = 0.362). Abbreviations: COPD = chronic obstructive pulmonary disease.

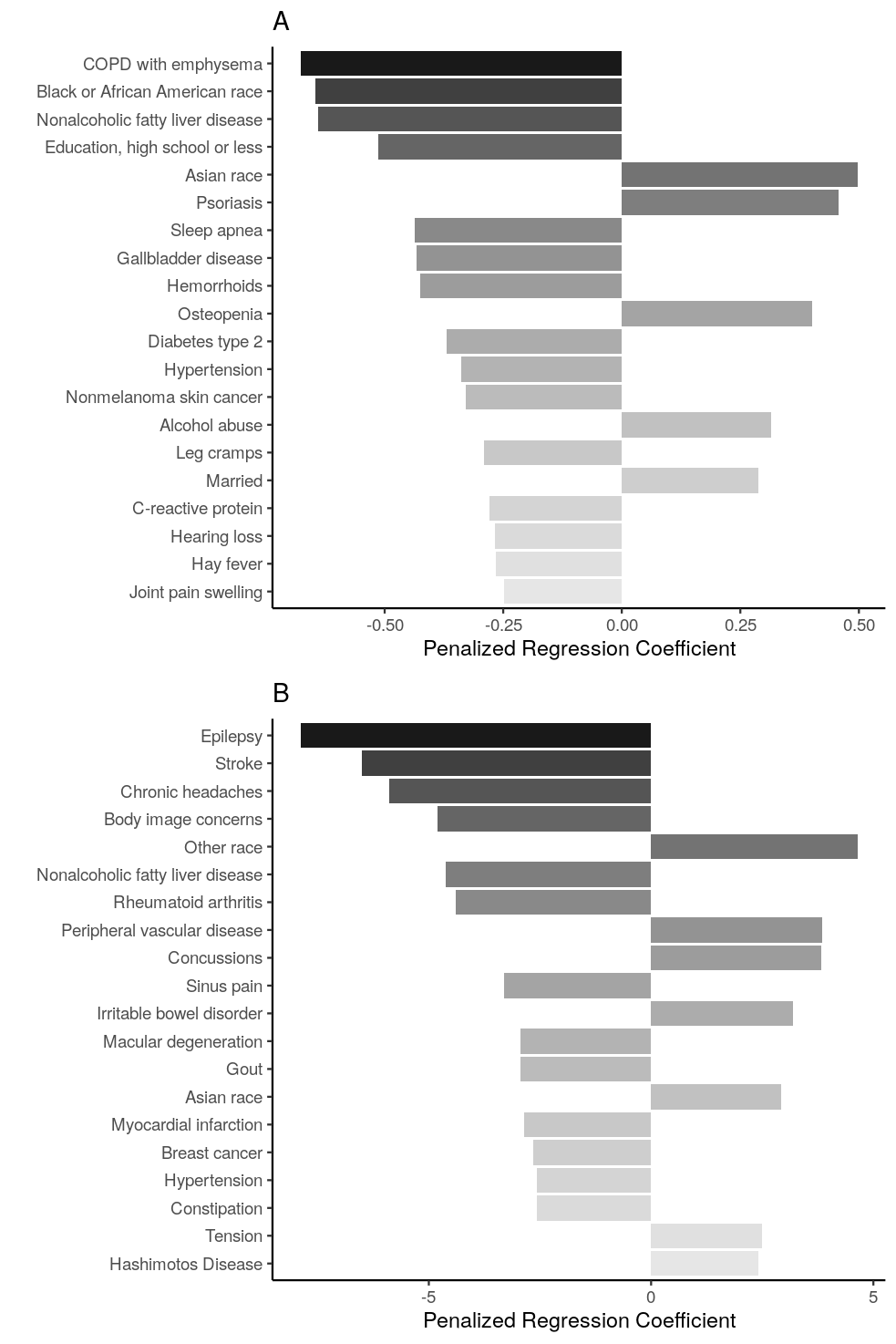

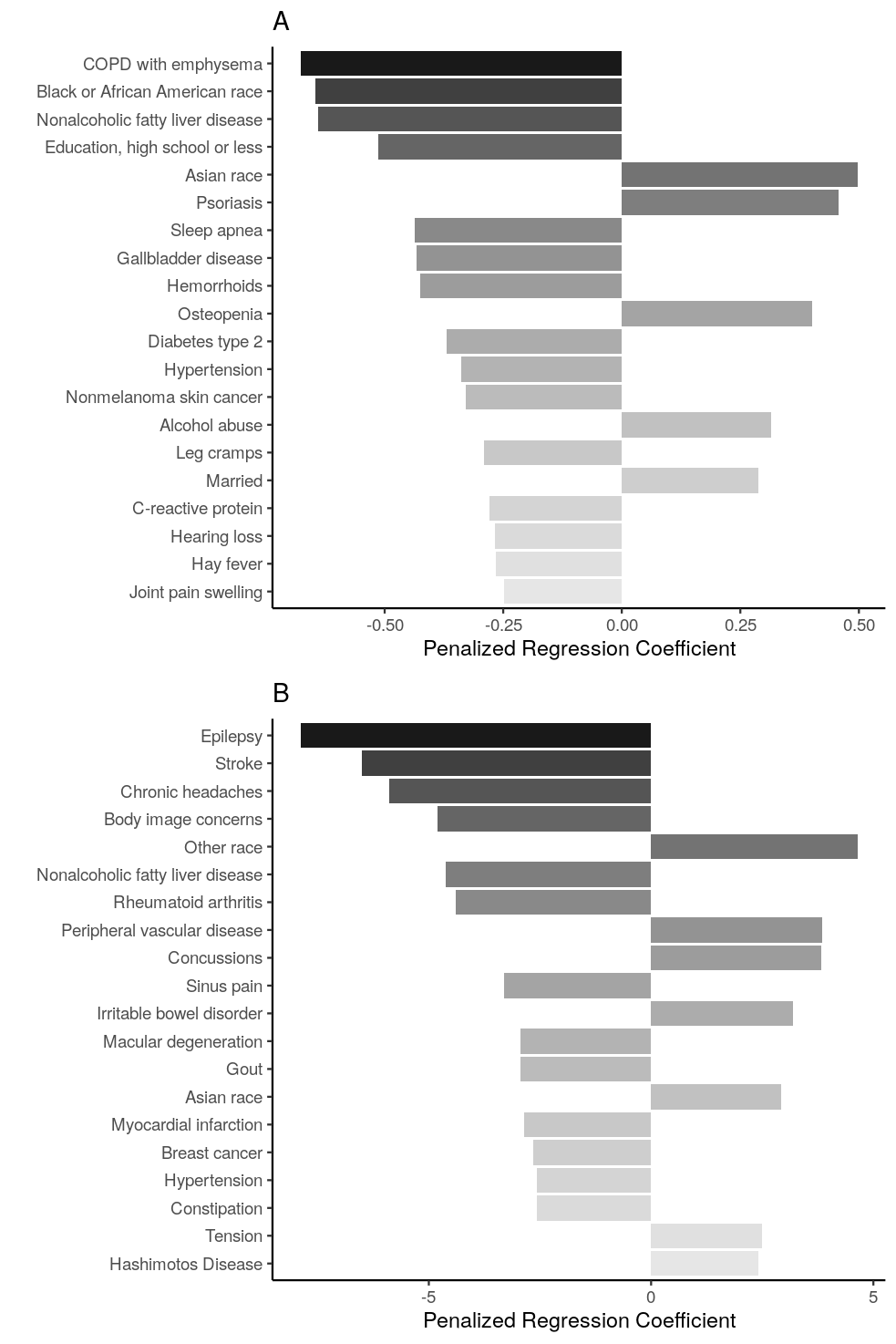

**Figure S2.** Top 20 regression coefficients of features selected from sENET regression model for sitting rising score, with the addition of features related to symptoms and allergies. Panel A: participants <65 years of age (ɑ = 1, λ= 0.0204); Panel B: participants ≥65 years of age (ɑ = 1, λ = 0.0413).

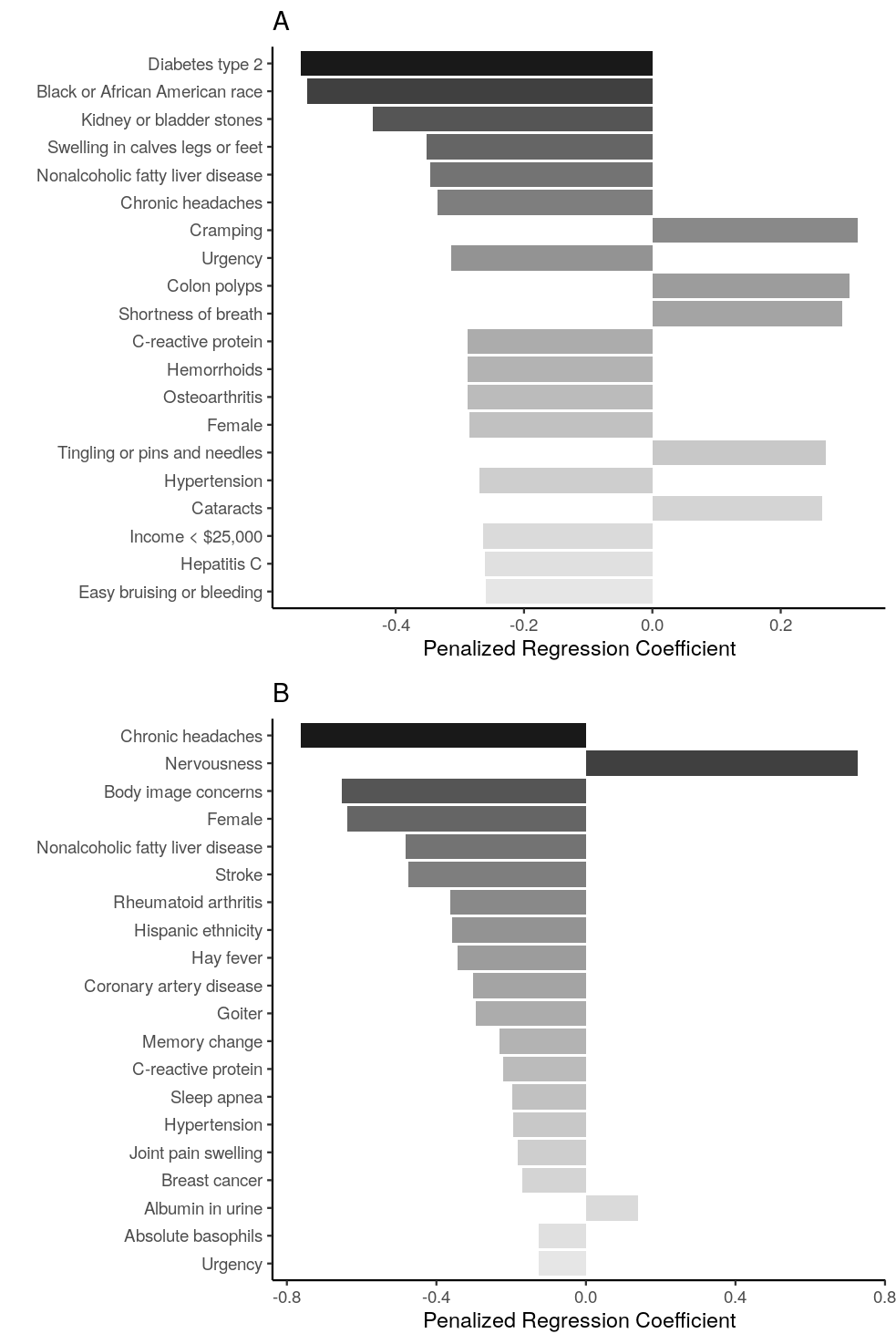

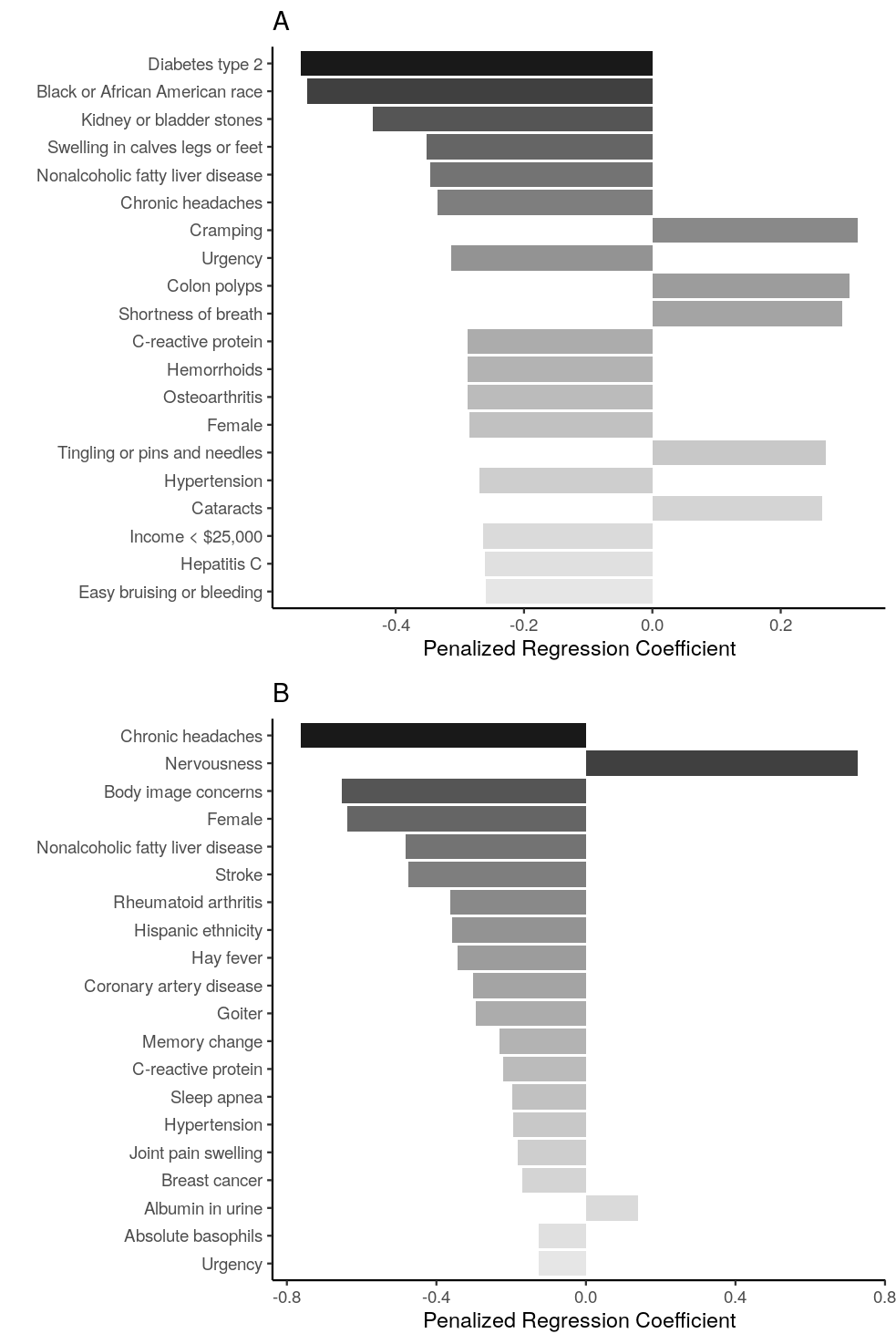

**Figure S3.** Top 20 regression coefficients of features selected from sENET regression model for 30-second chair stand test, with the addition of features related to symptoms and allergies. Panel A: participants <65 years of age (ɑ = 0.5, λ = 0.0294); Panel B: participants ≥65 years of age (ɑ = 0.5, λ = 0.108). Abbreviations: GERD = gastroesophageal reflux disease; GFR MDRD = glomerular filtration rate, Modification of DIet in Renal Disease equation; PTSD = post-traumatic stress disorder.

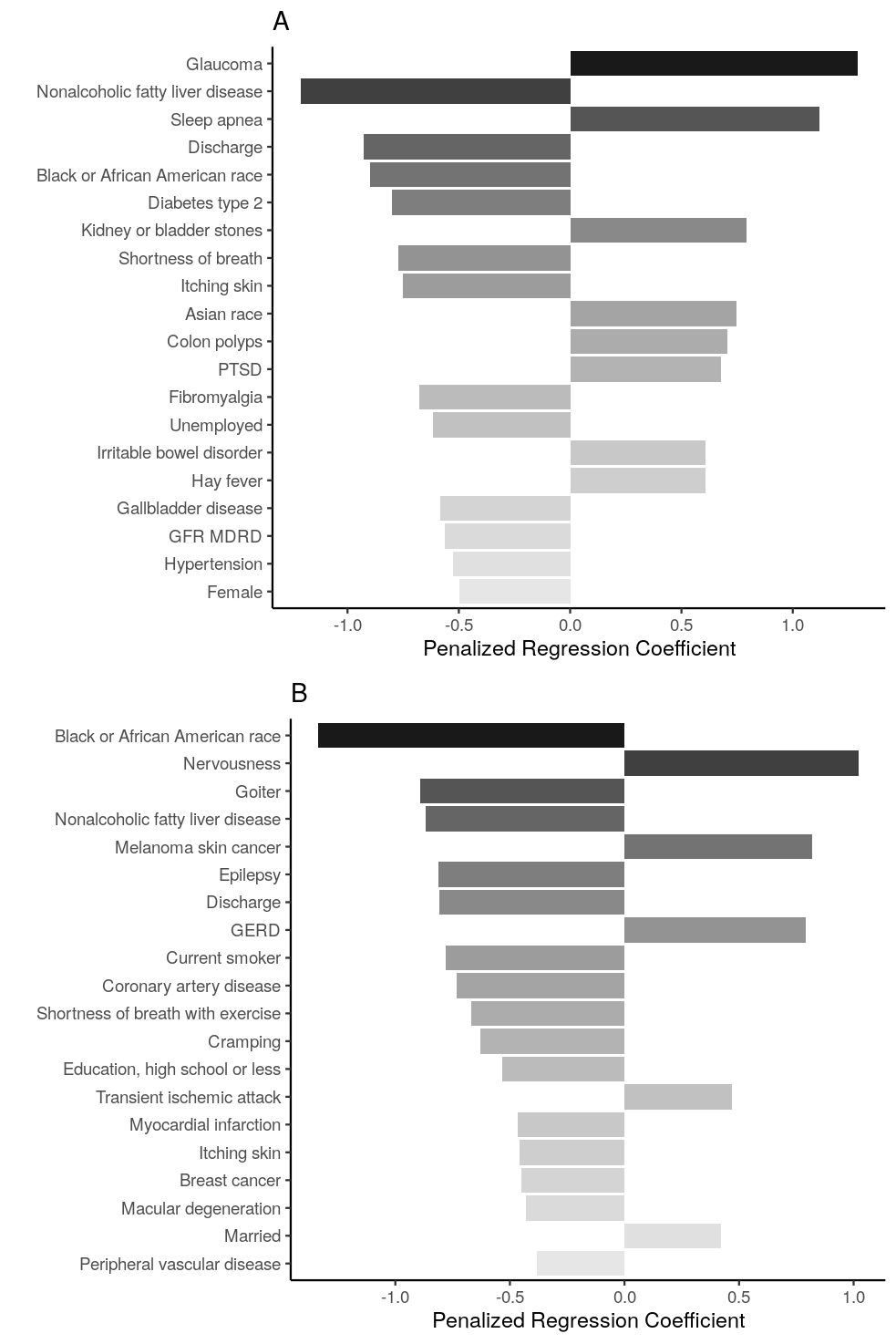

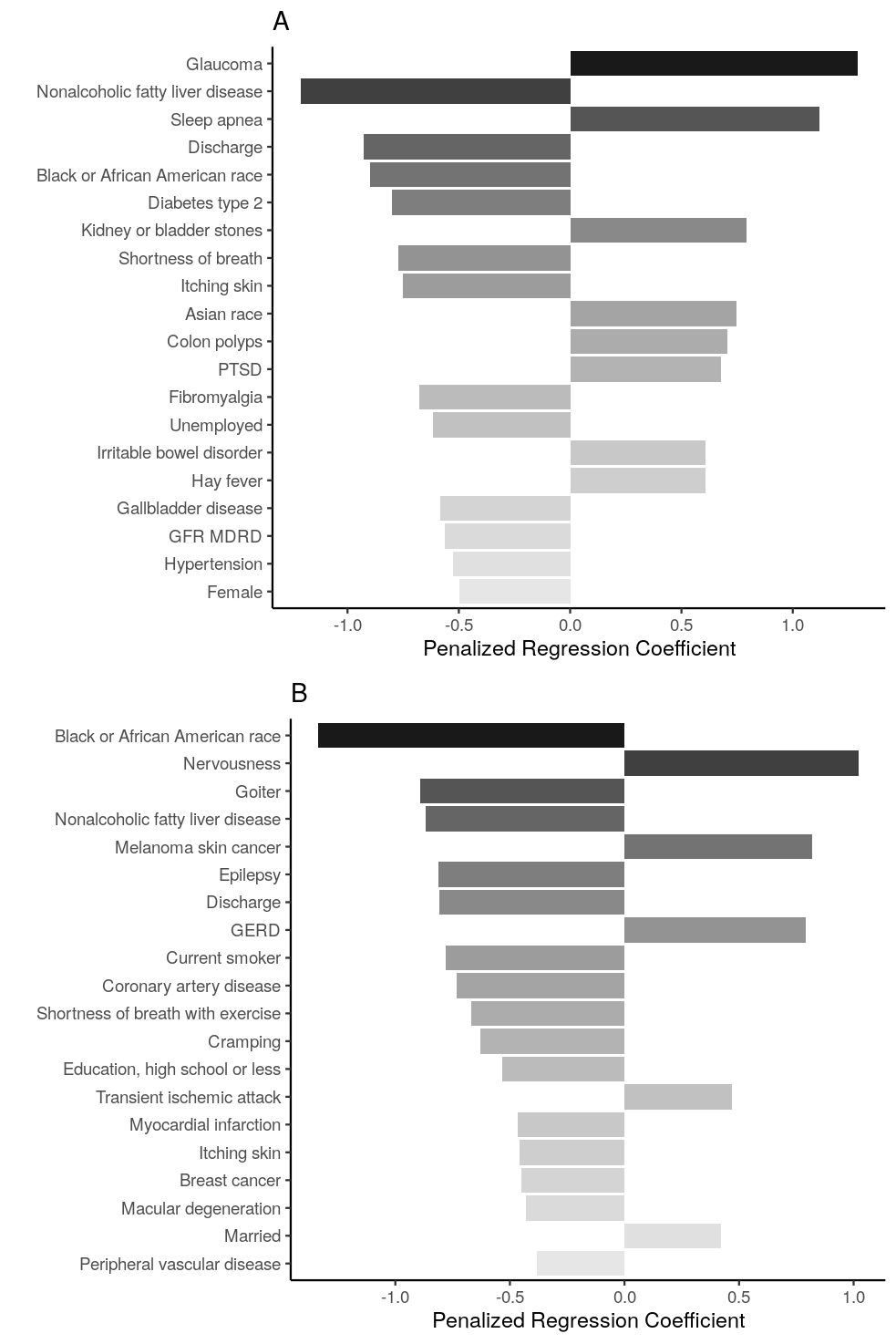
